# Development of a machine learning model to predict short duration HCV treatment response

**DOI:** 10.1101/2025.09.19.25336147

**Authors:** Joanne M Carson, Sebastiano Barbieri, Andrey Verich, Elise Tu, Andrew Lloyd, Gregory J Dore, Gail V Matthews, Marianne Martinello

**Affiliations:** Kirby Institute, University of New South Wales, Sydney, Australia; Queensland Digital Health Centre, University of Queensland, Brisbane, Australia; Centre for Big Data Research in Health, University of New South Wales, Sydney, Australia

**Keywords:** HCV, DAA, machine learning, artificial intelligence, modelling, treatment failure, personalised medicine

## Abstract

**Background:** Standard durations of direct acting antivirals (DAAs; 8–12 weeks) can be a barrier to HCV treatment initiation and completion among marginalised populations. This study developed a machine learning model to predict short-duration (4–6 weeks) DAA response using baseline clinical factors with potential to improve treatment uptake, cost-effectiveness and health system efficiency.

**Methods:** Baseline data from several short-duration DAA clinical trials and treatment discontinuations from real-world cohort studies were used. Multiple machine learning models were evaluated. Nested cross-validation was employed to optimise model hyperparameters and assess performance. Clinical utility was evaluated using Area Under Receiver Operator Characteristics (AUROC), Area Under Precision Recall Curve (AUPRC) and Matthews Correlation Coefficient (MCC). Threshold optimisation strategies were applied to balance model accuracy and DAA costs. Statistical analyses were conducted to estimate HCV RNA cutoffs predictive of treatment failure.

**Results:** Of 264 receiving short-duration DAAs (median 42 days; interquartile range 28-42), 94 (36%) experienced treatment failure. Predictors of failure included shorter durations, higher HCV RNA, higher AST–ALT ratio, genotype 3, and DAA class. The Elastic Net (regularised logistic regression) model demonstrated strong performance (AUROC: 83%; AUPRC: 73%). The Youden Index threshold balanced sensitivity (81%) and specificity (76%) with MCC of 0.56. A cost-optimized threshold, prioritizing retreatment minimization, achieved high sensitivity (98%) but reduced specificity (51%). HCV RNA cutoffs predictive of treatment failure were higher for protease+NS5A vs. NS5A+NS5B inhibitors.

**Conclusion:** Predictive models using baseline clinical data can identify individuals likely to respond to short-duration DAAs, with tailored thresholds enhancing clinical utility. Such models, if validated in larger datasets could facilitate HCV elimination efforts by improving treatment uptake, particularly for people who inject drugs, are homeless or incarcerated.

## Introduction

Despite the remarkable efficacy of direct-acting antivirals (DAAs) for hepatitis C virus (HCV), expanding treatment access among socially marginalized populations and dynamic populations remains a major challenge (1). Achieving HCV elimination requires increasing treatment coverage among key populations, such as people who inject drugs and people in prison, with heightened risk of HCV transmission.

While point-of-care testing has facilitated HCV diagnosis and rapid treatment initiation, the standard 8–12-week DAA regimens present barriers to treatment adherence and completion in vulnerable populations (2–4). People who inject drugs often face profound structural challenges, including unstable housing, homelessness, stigma and discrimination which may disrupt treatment (5). Similarly, people in prison are dynamic populations with high turnover rates, presenting challenges for prison health services when initiating treatment and for treatment completion (6).

Predictive models have the potential to personalize HCV treatments based on individual patient characteristics, thereby improving treatment uptake, adherence, cost-effectiveness, and overall health system efficiency (7–10). However, existing models predicting responses to short-duration regimens are limited by small sample sizes (30–95 participants), dependence on dynamic modelling frameworks, and the need for frequent on-treatment blood tests (11–16). These constraints make their implementation in real-world clinical settings challenging.

There is an urgent need for clinically applicable prediction tools that leverage baseline patient characteristics to forecast responses to shorter (4–6 week) DAA regimens. This analysis aimed to develop a machine learning model to predict sustained virologic response (SVR; cure) with short-duration DAA therapy, offering a potential pathway to optimize treatment for key populations.

## Methods

### Study population

This analysis included baseline data from adults (≥18 years) initiating short-duration DAA treatment for HCV in clinical trials (n=174; REACT, TARGETED, STRIVE4 (17–20)) or discontinued standard-duration treatment in real-world cohort or clinical trial studies (n=90; REACH-C, CEASE, STOP-C, SMART-C (21–24); **Supplementary Figure 1**). Eligible treatment durations ranged from 14 to 42 days for any DAA. For DAAs with a minimum recommended duration of 84 days, durations of 43 to 56 days (13% of the total sample) were included to increase the sample size and the ability of the model to detect patterns predictive of treatment response. Demographic, clinical, and treatment factors were collected at baseline in clinical trials or before treatment initiation in cohort studies. Treatment adherence (duration) was assessed systematically in trials and self- or health provider reported in real-world cohorts. Further details on the design and ethical approvals of included studies are available in **Supplementary Material**.

### Primary outcome

Sustained virologic response (SVR), defined as plasma HCV RNA below the limit of quantification 12 or more weeks after completing DAA treatment. Participants with reinfection at SVR confirmed by genotyping or sequencing, were classified as having achieved SVR (cure).

### Predictor candidates

Demographic factors (age, sex, race), clinical factors (baseline HCV RNA [log_10_ IU/mL], HCV genotype [GT], alanine aminotransferase [ALT], aspartate aminotransferase [AST], platelet count, liver stiffness indicative of fibrosis [transient elastography, kPa]), and treatment factors (DAA regimen and duration). Treatment duration, modelled as a continuous variable, was predominantly 28, 42, or 56 days (89%) due to clinical trial designs and standard pharmacy packaging.

### Data processing

Minimal data processing was undertaken to facilitate interpretability of machine learning outputs. DAA regimens were grouped into three classes - NS3/4A protease inhibitors (PI), NS5A inhibitors, and NS5B RNA-dependent RNA-polymerase inhibitors - to mitigate overfitting while retaining pharmacokinetic properties (25). AST-ALT ratio and APRI (AST-to-PLT Ratio Index) were calculated as proxy markers of liver inflammation, synthetic function and hepatocellular damage to provide clinically relevant features for model interpretation (26). Missing liver enzyme and stiffness data (<5%) were imputed using multiple imputation by chained equations (MICE). Transient elastography measures >12.5 kPa were considered diagnostic of cirrhosis.

### Model development

Standard logistic Regression, Elastic Net (regularised logistic regression combining L1 and L2 penalties), Support Vector Machine (SVM), Random Forest, AdaBoost, and XGBoost machine learning classifiers were developed (27). Nested cross-validation was used for hyperparameter optimization and performance evaluation. This framework separates hyperparameter tuning (inner folds) from performance assessment (outer folds) to reduce overfitting, enhance robustness, and improve model generalizability (28). Specifically, data were divided into randomized training and testing datasets using 3 × 5-fold nested cross-validation. During iterations of this procedure, the inner cross-validation folds were used to optimize hyperparameters. The final testing scores were computed exclusively on the outer folds, ensuring that the evaluation remained unbiased and independent of the hyperparameter tuning process. Class imbalance was addressed using Borderline-SMOTE (Synthetic Minority Oversampling Technique), which generates synthetic samples for the minority class near decision boundaries by interpolating between borderline instances and their nearest neighbours (29,30).

### Model evaluation

Models were evaluated based on performance metrics and clinical utility, emphasizing simplicity. Metrics computed for each fold and summarized as means with standard deviations (SD) included accuracy, sensitivity (true positive rate), specificity (true negative rate), precision (positive predictive value; PPV), negative predictive value (NPV), area under the receiver operating characteristic curve (AUROC), and area under the precision-recall curve (AUPRC) (31). AUROC measured model discrimination across thresholds, while AUPRC measured reliability in distinguishing treatment failures within imbalanced data. Matthews Correlation Coefficient (MCC) was included to evaluates prediction quality by balancing true and false outcomes, with scores ranging from −1 (total disagreement) to 1 (perfect prediction). Fowlkes-Mallows Index (FMI) was included to measures the balance between correctly predicted positive cases and avoiding false negatives, with scores ranging from 0 (poor) to 1 (perfect). As PPV, NPV, AUPRC and FMI depend on response prevalence in the test data, these metrics were used for model comparison but may not reflect real-world performance. SHAP (SHapley Additive exPlanations) values were computed to quantify feature importance, visualizing the magnitude and direction of each feature’s contribution to the predicted response.

### Model Calibration

Brier scores and calibration curves were computed on outer test folds of the nested cross-validation, with scores <0.2 considered indicative of good calibration. Predictions were interpolated onto a fixed probability grid and smoothed using the Savitzky–Golay filter to reduce noise while preserving trends, allowing visualization of agreement between predicted and observed event rates. An overconfidence penalty was applied to quantify miscalibration for highly confident but incorrect predictions (e.g., probabilities >85% assigned to negative outcomes or <15% to positive outcomes). This penalty, weighted twice that of other misclassification errors, was quantified as a weighted Brier score.

### Model threshold optimisation

Four thresholding strategies were employed to evaluate model performance: (1) Standard: default threshold of 0.5 classified probabilities ≥0.5 as positive and <0.5 as negative, assuming equal weight for false positives and false negatives. (2) Youden Index: threshold optimising the balance between sensitivity and specificity. (3) Treatment Cost Function: threshold minimizing a quadratic cost function that penalized overconfident errors with weights reflecting clinical priorities such as avoiding retreatment (4) Brier-Optimized: threshold minimizing Brier score emphasizing calibration between predicted and observed outcomes. Details on threshold optimization and performance metrics are available in the **Supplementary Material**.

### Statistical Analysis

Pearson’s correlation coefficient (r) was used to assess relationships between liver disease markers, visualized with scatter plots. Differences in baseline HCV RNA levels and liver disease markers between acute and chronic HCV phases were analysed using the Mann-Whitney U test and visualized with violin plots. Logistic regression was used to estimate adjusted odds ratios (AORs) for predictors of treatment failure. Variance Inflation Factor (VIF) >0.5 and Phi coefficient (|φ|) >±0.5 were considered indicative of collinearity. HCV RNA cut-off values predictive of treatment failure for different treatment durations and GT combinations were estimated using the machine learning model. To do this, HCV RNA values were generated as a linearly spaced array that ranged the upper and lower limits of HCV RNA quantification (1-8 log_10_ IU/L), with relevant factors activated and others deactivated or set to the mean value. Predicted probabilities of treatment failure for HCV RNA values were plotted with 95% confidence intervals (CIs) computed via bootstrapping (n=1000 samples with replacement).

A flow chart illustrating stages of model development is presented in **Supplementary Figure 2**. This analysis follows the Transparent Reporting of a Multivariable Prediction Model for Individual Prognosis or Diagnosis (TRIPOD) guidelines (**Supplementary Table 1**(31)). Analyses were performed in Python version 3.12.3. Model code is available at https://github.com/jojocarson/hcv_tx_response

## Results

### Population Characteristics

Of 264 individuals treated for HCV with short-duration DAAs, 170 (64%) achieved SVR and 94 (36%) experienced treatment failure. Median age of the study populations was 43 years (IQR 32–52); most were male (93%), Caucasian/White (85%), and enrolled in clinical trials (69%; **Table 1**). Characteristics of the clinical trial and cohort populations are displayed in **Supplementary Table 2**. Recent injecting drug use (44%), identifying as a gay or bisexual man (58%), and HIV coinfection (50%) were common, whereas current incarceration was less common (14%). Median baseline HCV RNA was 5.9 log_10_ IU/mL (IQR 5.1–6.6), lower in the SVR group compared to the treatment failure group (5.6 vs. 6.3 log_10_ IU/mL). HCV GT1 was most prevalent (63%), occurring more frequently in those with SVR compared to treatment failure (72% vs. 47%), whereas GT3 (25%) occurred more frequently in the treatment failure group (16% vs. 43%). Acute HCV (32%: average estimated infection duration 123 days [SD 36]) occurred more frequently in the SVR compared to failure group (43% vs. 11%). While HCV RNA levels were comparable between acute and chronic infections (p=0.314), AST-ALT ratio in acute infections was lower (p<0.001; **Supplementary Figure 3**).

**Table 1.** Characteristics of the study population stratified by short-duration HCV treatment response.

|  | SVR<br>(n=170) | Treatment<br>Failure<br>(n=94) | Total<br>(n=264) |
| --- | --- | --- | --- |
| Age, median [Q1, Q3] | 42 [32, 50] | 44 [31, 55] | 43 [32, 52] |
| Male gender, n (%) | 161 (94.7) | 84 (89.4) | 245 (92.8) |
| Caucasian/white race, n (%) | 137 (80.6) | 86 (91.5) | 223 (84.5) |
| Gay or bisexual man, n (%) | 133 (78.2) | 20 (21.3) | 153 (58.0) |
| IDU past six months, n (%) | 72 (43.9) | 32 (43.2) | 104 (43.7) |
| Incarcerated, n (%) | 8 (4.7) | 30 (31.9) | 38 (14.4) |
| HIV, n (%) | 112 (65.9) | 19 (20.2) | 131 (49.6) |
| Clinical trial, n (%) | 155 (91.2) | 28 (29.8) | 183 (69.3) |
| Acute HCV, n (%) | 74 (43.5) | 10 (10.6) | 84 (31.8) |
| HCV RNA log<sub>10</sub> IU/L, median [Q1, Q3] | 5.6 [4.9, 6.3] | 6.3 [5.5, 6.8] | 5.9 [5.1, 6.6] |
| Genotype, n (%) |  |  |  |
| 1 | 123 (72.4) | 44 (46.8) | 167 (63.3) |
| 3 | 27 (15.9) | 40 (42.6) | 67 (25.4) |
| 2, 4 or 6 | 20 (11.7) | 10 (10.6) | 30 (11.3) |
| ALT, median [Q1, Q3] | 136 [68, 289] | 82 [46, 151] | 110 [55, 237] |
| AST, median [Q1, Q3] | 64 [38, 125] | 55 [40, 81] | 62 [38, 104] |
| PLT, median [Q1, Q3] | 235 [200, 271] | 228 [181, 268] | 233 [197, 271] |
| AST-ALT ratio, median [Q1, Q3] | 0.5 [0.4, 0.7] | 0.7 [0.5, 1.0] | 0.5 [0.4, 0.8] |
| APRI, median [Q1, Q3] | 0.7 [0.4, 1.5] | 0.6 [0.4, 1.0] | 0.7 [0.4, 1.3] |
| Liver stiffness (kPa), median [Q1, Q3] | 6.0 [4.9, 7.4] | 6.4 [4.9, 8.1] | 6.1 [4.9, 7.7] |
| Cirrhosis, n (%) | 9 (5.3) | 6 (6.4) | 15 (5.7) |
| DAA classes, n (%) |  |  |  |
| NS5A+NS5B<sup>1</sup> | 90 (52.9) | 72 (76.6) | 162 (61.4) |
| PI+NS5A<sup>2</sup> | 50 (29.4) | 20 (21.3) | 70 (26.5) |
| PI+NS5A+NS5B<sup>3</sup> | 30 (17.6) | 2 (2.1) | 32 (12.1) |
| Duration (days), median [Q1, Q3] | 42 [42, 42] | 35 [28, 42] | 42 [28, 42] |

**Abbreviations:** IDU, injecting drug use; ALT, alanine aminotransferase; AST, aspartate aminotransferase; PLT, platelet count; APRI, AST to Platelet Ratio Index, DAA, direct acting antiviral; PI, protease inhibitor; SVR, sustained virologic response
<sup>1</sup>NS5A+NS5B inhibitors: sofosbuvir-velpatasvir (n=134); sofosbuvir-ledipasvir (n=17); sofosbuvir+daclatasvir (n=11), <sup>2</sup>PI+NS5A inhibitors: glecaprevir-pibrentasvir (n=70); <sup>3</sup>PI+NS5A+NS5B inhibitors: ombitasvir-paritaprevir-ritonavir+dasabuvir (n=30); glecaprevir-pibrentasvir+sofosbuvir (n=1); grazoprevir-elbasvir+sofosbuvir (n=1)

Despite differences in AST-ALT ratio by HCV phase, AST-ALT ratio (0.5 vs. 0.7), APRI (0.7 vs. 0.6), and liver fibrosis (6.0 vs. 6.4 kPa) scores were similar between SVR and failure groups. Cirrhosis was uncommon, occurring in only 6% of participants. ALT and AST were strongly correlated (r=0.87), reflecting consistent elevations in both liver enzymes. APRI was moderately correlated with fibrosis score (r=0.54), whereas AST-ALT ratio showed negligible to weak correlations with APRI (r=-0.06) and fibrosis scores (r=0.14; **Supplementary Figure 4)**. DAA classes used to treat HCV in short durations were NS5A+NS5B (61%), PI+NS5A (27%), and PI+NS5A+NS5B (12%) inhibitor combinations. Treatment durations ranged from 14-56 days, with 28-day (23%), 42-day (55%), and 56-day (11%) durations most common.

### Logistic Regression Model

Higher AST-ALT ratio (AOR 7.48; 95% CI 2.50–22.39; p<0.001), higher baseline HCV RNA (AOR 2.10 per log_10_ IU/L; 95% CI 1.51–2.92; p<0.001), and GT3 infection (AOR 3.16; 95% CI 1.52–6.61; p=0.002) were associated with increased likelihood of treatment failure **(Supplementary Table 5)**. PI+NS5A inhibitors (vs. NS5A+NS5B inhibitors; AOR 0.31; 95% CI 0.14– 0.68; p=0.004) and longer treatment durations (AOR 0.90 per day; 95% CI 0.86–0.94; p<0.001) were associated with SVR. age, sex, race, APRI, and liver fibrosis were not significant predictors. Acute HCV, HIV coinfection, clinical trial participation, and identifying as a gay or bisexual man exhibited high levels of collinearity (|φ|>±0.7) and were excluded from the analysis. The standard logistic regression model, restricted to significant predictors, demonstrated good performance in nested cross-validation (AUROC 0.78 [SD 0.04]; AUPRC 0.67 [SD 0.08]; **Supplementary Figure 5**). Logistic regression calibration (Brier score: 0.20 [SD 0.03]), worsened slightly (0.22 [SD 0.04]) when the penalty for highly confident but incorrect predictions was applied.

### Elastic Net Model

Building on significant predictors identified in logistic regression, the Elastic Net (regularized logistic regression) model demonstrated superior performance in nested cross-validation (AUROC 0.83 [SD 0.05]; AUPRC 0.73 [SD 0.08]; **Figure 1**). At the default threshold, the Elastic Net model achieved higher accuracy (0.74 vs. 0.69), FMI (0.70 vs. 0.64) and MCC (0.49 vs. 0.39; **Table 2**) compared to logistic regression. The Elastic Net model showed good calibration (Brier score: 0.18 [SD 0.02]), which worsened slightly (0.21 [SD 0.03]) when overconfident predictions were penalised. Of note, most overconfident predictions were SVR misclassified as treatment failure (6/7); were NS5A/NS5B inhibitor treated (6/7), GT1 infections (5/7), and from real-world cohorts (4/7).

**Table 2.** Performance metrics of Logistic Regression and Elastic Net models across thresholding strategies. Performance metrics are presented as mean values with standard deviations, derived from nested cross-validation. Metrics for the Elastic Net model are displayed for default and optimized thresholds (Youden Index, Quadratic, and Brier score).

|  | Logistic | Elastic Net | Elastic Net Optimised Thresholds |  |  |
| --- | --- | --- | --- | --- | --- |
| Metric, (Mean, [SD]) | Default | Default | Youden Index | Quadratic | Brier Score |
| Sensitivity | 0.76 [0.11] | 0.83 [ 0.07] | 0.81 [0.09] | 0.98 [0.03] | 0.65 [0.11] |
| Specificity | 0.64 [0.07] | 0.68 [0.04] | 0.76 [0.09] | 0.51 [0.10] | 0.87 [0.07] |
| PPV | 0.54 [0.07] | 0.59 [0.04] | 0.67 [0.07] | 0.53 [0.03] | 0.75 [0.09] |
| NPV | 0.83 [0.06] | 0.88 [0.04] | 0.88 [0.05] | 0.98 [0.02] | 0.83 [0.06] |
| Accuracy | 0.68 [0.06] | 0.74 [ 0.03] | 0.78 [0.05] | 0.68 [0.05] | 0.79 [0.04] |
| F1 Score | 0.63 [0.08] | 0.69 [0.04] | 0.72 [0.05] | 0.69 [0.02] | 0.68 [0.09] |
| FMI | 0.64 [0.08] | 0.70 [0.05] | 0.73 [0.06] | 0.72 [0.02] | 0.69 [0.08] |
| MCC | 0.39 [0.13] | 0.49 [0.07] | 0.56 [0.08] | 0.50 [0.05] | 0.55 [0.09] |

**Abbreviations:** PPV, positive predictive value; NPV, negative predictive value; FMI, Fowlkes-Mallows Index; MCC, Matthews Correlation Coefficient

**Figure 1.**
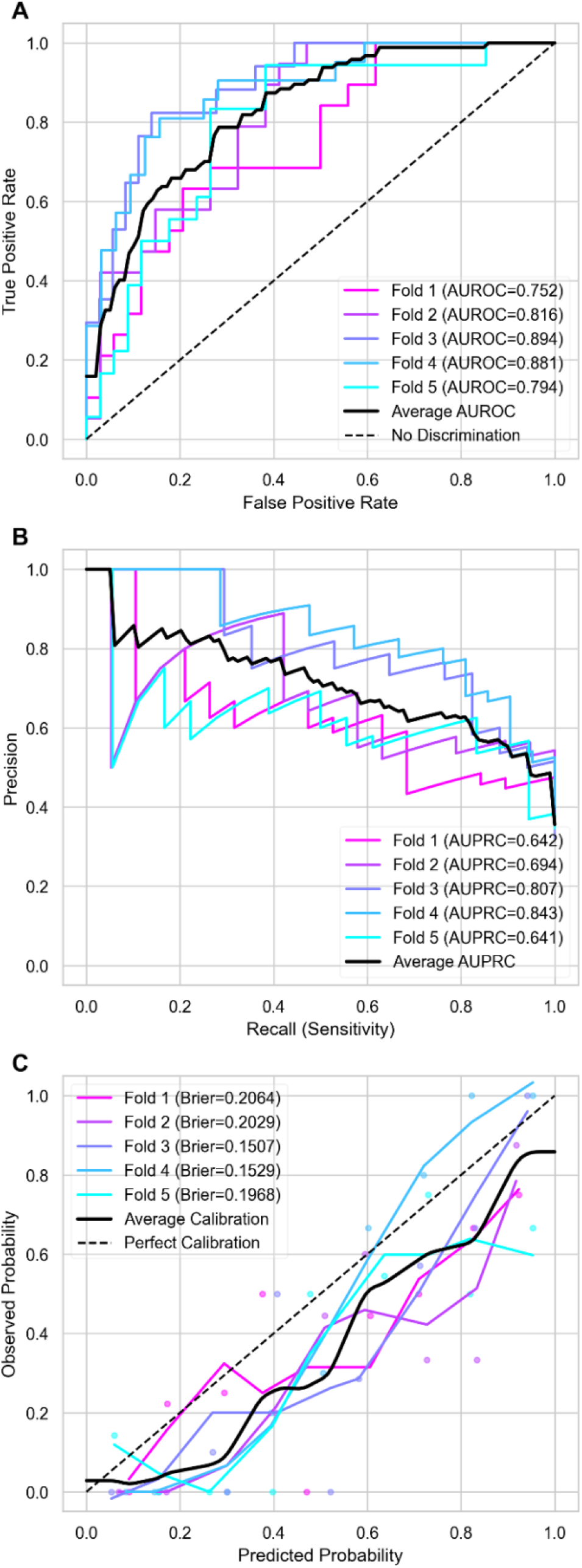
Nested Cross-Validation for Elastic Net Model Performance. Evaluation of the Elastic Net model’s predictive accuracy through nested cross-validation, presented with **(A)** AUROC (Area Under the Receiver Operating Characteristic Curve), **(B)** AUPRC (Area Under the Precision-Recall Curve), and **(C)** Brier Calibration Curve to provide insight with Savitzky–Golay smoothing as to how well the model generalizes across varying subsets of data for predicting HCV treatment response.

Optimized thresholds were used to enhance the Elastic Net model utility. The Youden Index achieved the best balance between sensitivity (0.81) and specificity (0.76), with the highest F1 score (0.72), FMI (0.73) and MCC (0.56; **Table 2**). The quadratic threshold, designed to minimize retreatment costs while penalizing overly confident but incorrect classifications, prioritized sensitivity (0.98) at the expense of specificity (0.51) but maintained a comparable FMI (0.72). Similarly, the Brier Score threshold achieved high specificity (0.87) at the expense of sensitivity (0.65) while maintaining comparable MCC (0.55).

SHAP values were computed to assess the relative importance of features in the Elastic Net model. Treatment duration, baseline HCV RNA, and AST–ALT ratio exerted the greatest influence on predictions of treatment response (**Figure 2**). Shorter DAA durations, higher HCV RNA levels, higher AST-ALT ratios, and genotype 3 infection were predictive of treatment failure. PI+NS5A inhibitor combinations were predictive of SVR, whereas NS5A+NS5B inhibitor combinations were predictive of treatment failure.

**Figure 2.**
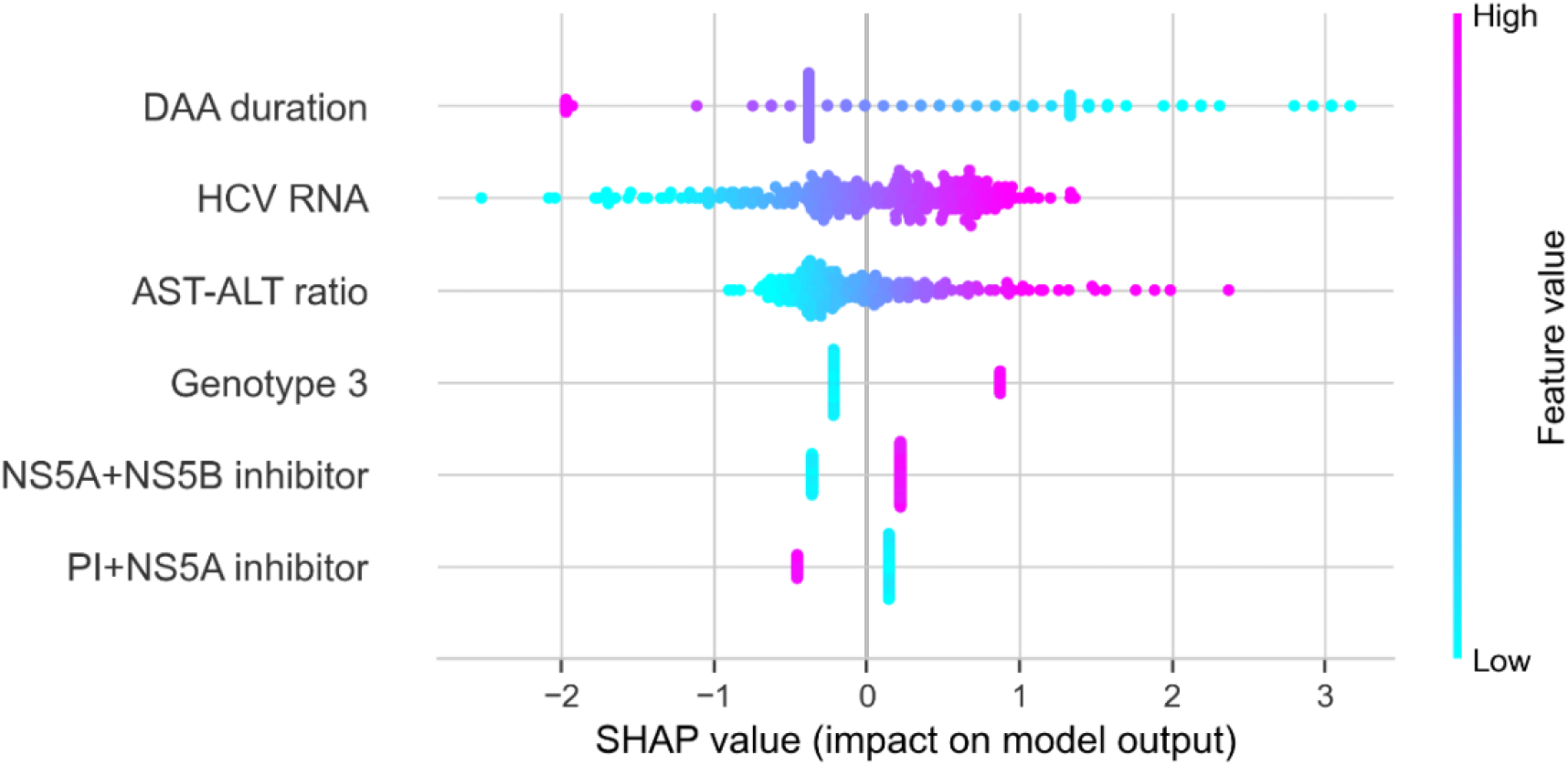
Elastic Net Feature importance scores. SHAP (SHapley Additive exPlanations) plot illustrating feature contributions to the Elastic Net model’s predictions of treatment failure in HCV patients. Each point represents a SHAP value for a feature in a given observation, where colour indicates feature value (e.g., high or low). Positive SHAP values suggest a higher probability of treatment failure, while negative values suggest a greater likelihood of SVR. **Abbreviations:** DAA, direct acting antiviral; HCV, hepatitis C virus; ALT, alanine aminotransferase; AST, aspartate aminotransferase; PI, protease inhibitor; SVR, sustained virologic response

Baseline HCV RNA cut-offs used by the Elastic Net model to classify treatment failure (at default 0.5 threshold) were estimated for different combinations of DAA class, treatment duration, and genotype, with other variables fixed. For GT3, the estimated HCV RNA cut-off values were higher for PI+NS5A vs. NS5A+NS5B inhibitor treatment for 28-day durations (4.28 vs. 2.49 log_10_ IU/mL) and for 42-day durations (6.95 vs. 5.17 log_10_ IU/mL; **Figure 3**). Similarly, for non-GT3 infections, PI+NS5A inhibitors demonstrated higher HCV RNA cut-offs for treatment failure than NS5A+NS5B regimens for both 28-day (5.89 vs. 4.08 log_10_ IU/mL) and 42-day durations (8.54 vs. 6.78 log_10_ IU/mL).

**Figure 3.**
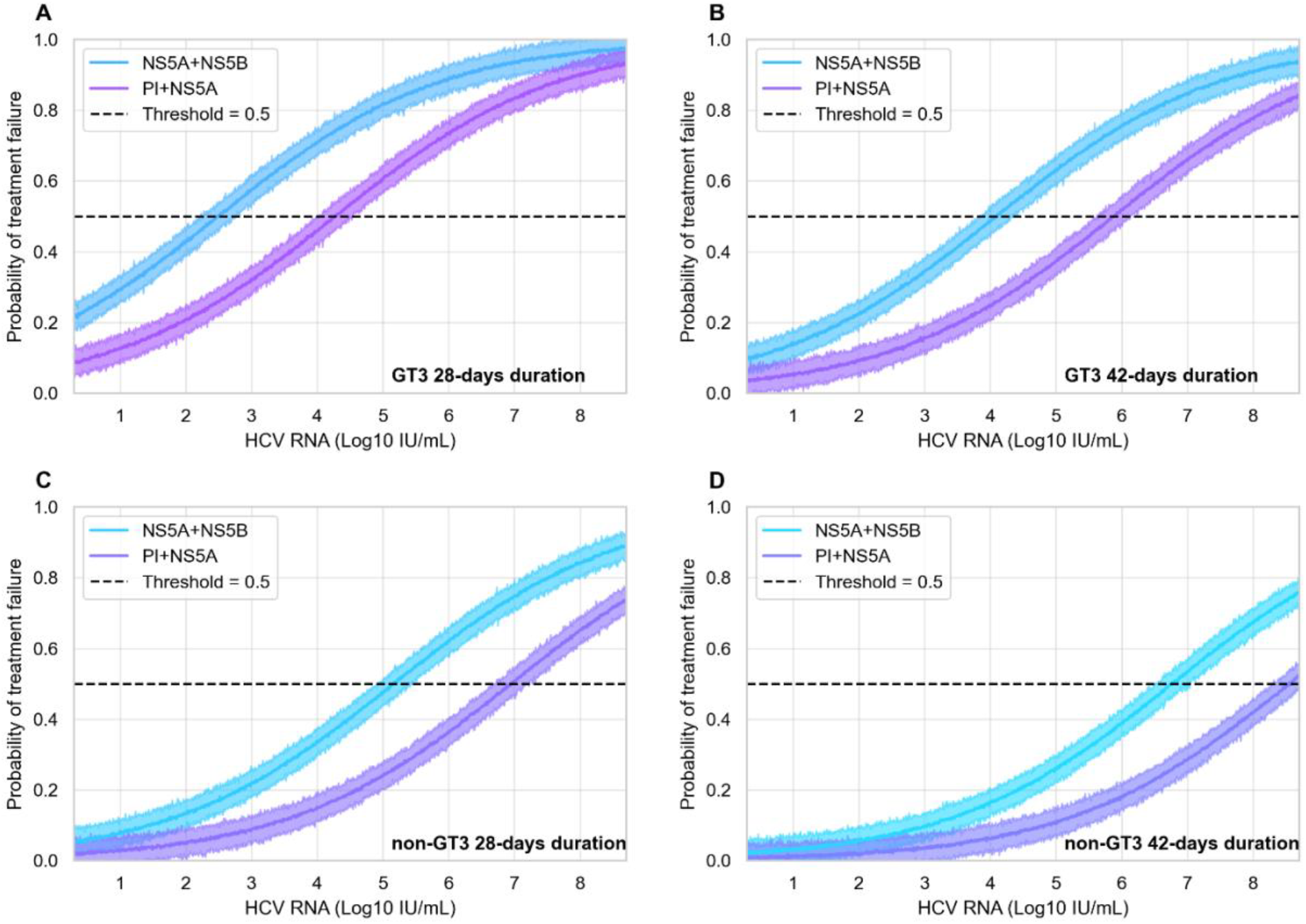
HCV RNA values predictive of treatment failure. Predicted probabilities of treatment failure are plotted against baseline HCV RNA levels, as estimated by the Elastic Net model, with bootstrapped 95% confidence intervals. The curves illustrate differences in HCV RNA cutoffs for predicting treatment response for NS5A+NS5B and PI+NS5A regimens, stratified by treatment duration and genotype. Results are shown for genotype 3 infections treated with **(A)** 28-day and **(B)** 42-day durations; and for non-genotype 3 infections treated **(C)** 28-day and **(D)** 42-day durations with. AST-ALT ratio was fixed at the mean value in all scenarios, and HCV RNA cutoff points correspond to probabilities crossing the default 0.5 threshold. **Abbreviations:** HCV, hepatitis C virus; GT, genotype; PI, protease inhibitor.

### XGBoost Model

Models incorporating a greater number of demographic, clinical, and treatment factors were developed for comparison. In nested cross-validation, XGBoost demonstrated similar performance to the simpler Elastic Net model (AUROC 0.82 [SD 0.04], AUPRC 0.71 [SD 0.08]; **Supplementary Figure 6**), with lower sensitivity (0.69 [SD 0.10] vs. 0.74 [SD 0.03]) but higher specificity (0.79 [SD 0.04] vs. 0.68 [SD 0.03]) at the default threshold. The XGBoost model initially demonstrated comparable calibration (Brier score: 0.18 [SD 0.02]); however, the application of the overconfidence penalty resulted in a worsened calibration (Brier score: 0.27 [SD 0.08]). This deterioration, characterized by a tendency toward overconfident yet incorrect classifications, may suggest worse generalizability, potentially indicative of overfitting. Consistent with the Logistic and Elastic Net Models the SHAP value analysis for the XGBoost model identified DAA duration, HCV RNA, and AST-ALT ratio as the most influential predictors **(Supplementary Figure 7)**.

## Discussion

We developed a machine learning model to predict response to short-duration HCV treatment using data from real-world cohorts and clinical trials. The study population included key groups with HCV transmission risk, such as people with recent injecting drug use and people in prison. The Elastic Net algorithm emerged as the most efficient predictive model, achieving strong performance (AUROC: 83%; AUPRC: 73%) with a small number of clinical predictors, including treatment duration, baseline HCV RNA, liver enzymes (AST-ALT ratio), genotype, and DAA class.

The Elastic Net model’s performance surpassed that of more complex models, demonstrating simpler, interpretable models using readily available clinical data to predict treatment response can be effective and may minimize overfitting. Tailored threshold optimization enhanced real-world utility: the Youden threshold provided balanced performance (sensitivity 81%; specificity 76%), the treatment cost-weighted threshold prioritized minimising retreatment (sensitivity 98%; specificity 51%). Alternatively, the Brier-optimized threshold, which emphasized calibration (specificity 87%; sensitivity 65%), may aid retreatment decisions by distinguishing treatment failure from reinfection when HCV viremia is detected following discontinuation of standard-duration HCV treatment.

The short-duration model outperformed a previous machine learning model for standard-duration treatment, largely due to greater specificity in predicting failure. The prior study, which utilised a large but imbalanced dataset (n=6,525; 5% treatment failure), also identified Elastic Net as the most parsimonious model and selected 27 predictors, including some overlaps such as treatment duration and liver function markers (32). However, key differences emerged: baseline HCV RNA, a strong predictor of short-duration response, was not predictive for standard-duration treatment. In our model, the influence of AST-ALT ratio likely reflected acute HCV infection (predictive of SVR), whereas in the prior study, elevated liver enzymes were likely correlated with advanced liver disease (predictive of failure). The previous cohort consisted exclusively of chronically infected, older individuals predominantly treated with genotype-specific DAAs, and identified several medications as influencing treatment response, many of which were linked to chronic comorbidities associated with advanced HCV.

In contrast, our study focused on a population more reflective of those with ongoing HCV transmission risk. A high proportion of participants had acute HCV infection (i.e., were recently infected), most had minimal liver fibrosis, and were treated with contemporary pangenotypic DAAs. This distinction is crucial, as it underscores the model’s applicability to populations where short-duration treatment could have significant contextual utility - such as people who inject drugs and those in prison – settings where increasing treatment access and reducing transmission are paramount.

HCV-RNA cut-offs predicting treatment failure were higher for PI+NS5A compared with NS5A+NS5B combinations. These thresholds for HCV RNA cut-offs can be used to guide standard-duration treatment choice; for patients at risk of treatment discontinuation, DAA regimens with faster viral suppression can maximise probability of SVR (16). PI+NS5A combinations (e.g., glecaprevir-pibrentasvir) inhibit replication rapidly because both drugs reach high intracellular levels, undergo little metabolism, and—in the case of pibrentasvir—have a long half-life contributing to sustained antiviral activity (25). In contrast, NS5A–NS5B regimens rely on sofosbuvir, a prodrug which must be phosphorylated and is rapidly converted to less active metabolites, consistent with the lower RNA thresholds that predicted failure in our model (25). These thresholds were lowest for genotype 3, a strain with faster replication that accelerates hepatocyte damage and structural variations in NS5A that weaken inhibitor binding affinity (33,34). Clarifying the minimal effective therapeutic duration may however help set target exposure windows for the long-acting injectable DAAs currently in development (35).

Our findings also enable a pragmatic algorithm to guide retreatment decisions when HCV viraemia is detected after discontinuation of a standard-duration DAA course, which is particularly useful among people who inject drugs or people in custodial settings, where treatment interruptions, delayed SVR testing, and uncertain reinfection status are a frequent reality (36). At re-presentation, the model could be applied to estimate probability of treatment failure or reinfection. Because the algorithm relies solely on routinely collected data, it can be embedded in electronic prescribing systems and used even when genotyping is unavailable or reinfection involves the same genotype, adding no laboratory cost and negligible clinician workload. This simplicity is key – predictive models that increase the complexity of care or health expenditure - are unlikely to be successfully implemented in practice (37).

Predictive models that increase the complexity of care, clinician workload, and medical expenditure - such as those requiring additional diagnostic tests or clinical assessment - are unlikely to be widely adopted (37). Because the model uses only routinely collected baseline clinical variables, it adds no laboratory cost and minimal clinician workload - features critical for successful implementation (37,38). Even so, with patients ranking probability of SVR above the convenience of shorter duration therapy when making treatment decisions (39), conventional 8–12 week courses remain the safest default, particularly for those at risk of non-adherence or disengagement with care (2,5,40).

A principal limitation of our analysis is the predominance of acute and recent HCV infection. In the interferon (IFN) era, acute HCV infection required markedly shorter treatment durations because exogenous IFN-α augmented innate–adaptive responses; chronic infection, characterised by T-cell exhaustion and fibrotic remodelling, required longer durations (41). DAAs act directly on viral proteins and are therefore less dependent on host immunity (25), however immune activity during acute infection may accelerate viral clearance and compensate for abbreviated dosing. Evidence to quantify this advantage is sparse; most reports of “short-duration” DAAs for chronic HCV have come from real-world cohorts where treatment was discontinued and adherence prior to discontinuation may be uncertain or suboptimal. Rigorous trials testing 4- to 6-week regimens in chronic HCV, followed by pragmatic studies with objective adherence monitoring, are needed to support personalised treatment programs.

Our study has other limitations which must be acknowledged. Although the model achieved moderate diagnostic accuracy (MCC 0.56 with Youden threshold), it has not been externally validated and therefore does not yet meet the high standards required for clinical translation. The data used to develop the model was small and lacked gender and racial diversity; only a few participants had advanced fibrosis, limiting generalisability. We used Borderline-SMOTE to offset class imbalance and nested cross-validation to avoid data leakage; however strong correlation between acute phase, HIV, gay or bisexual identity and clinical trial participation made it challenging to disentangle individual effects. This correlation may simply mark higher engagement in care, regular HCV screening, and better adherence. Laboratory values were obtained on the treatment-start day in clinical trials but may have been measured days or weeks earlier in real-world cohorts, potentially introducing measurement error. AST–ALT ratio, a proxy for acute HCV, can also be influenced chronic alcohol use or Metabolic Dysfunction-Associated Fatty Liver Disease (MELD) therefore should be interpreted with caution. Finally, we analysed drug classes rather than individual regimens, so intraclass pharmacokinetic differences may not have been captured.

Future studies should focus on validating our findings in larger, more diverse cohorts that are representative of populations likely to benefit from short-duration HCV treatment. Comprehensive cost-benefit analyses will be essential to evaluate the economic viability of implementing such models across various healthcare settings. Moreover, understanding both patient and prescriber perspectives and preferences are crucial for the successful translation of any predictive model into clinical practice.

In conclusion, this study shows that simple machine learning models using readily available clinical data can predict response to short-duration HCV treatment with good accuracy. However, larger, more diverse datasets are needed to ensure predictions of treatment response are robust and generalisable. Despite potential variation in clinical utility, such predictive models could aid HCV elimination efforts by supporting treatment expansion in dynamic populations.

## Data Availability

The datasets used in this study are held securely at the Kirby Institute, UNSW Sydney. Due to the sensitive nature of this data, it cannot be made publicly available however, data used in this analysis may be made available for research purposes upon reasonable request to the corresponding author. The code used to develop the models is publicly available at https://github.com/jojocarson/hcv_tx_response

## Declarations

### Ethics approval and consent to participate

Not applicable.

### Consent for publication

Not applicable.

### Author contributions

**Carson JM:** Conceptualization, Data curation, Data analysis, Methodology, Visualization, Writing – original draft, Writing – review & editing

**Barbieri S:** Conceptualization, Methodology, Supervision, Writing – review & editing

**Verich A:** Laboratory analysis, Writing – review and editing.

**Tu E:** Data curation, Writing – review and editing.

**Lloyd A:** Writing – review & editing

**Dore GJ:** Writing – review & editing

**Matthews GV:** Writing – review & editing

**Martinello M:** Conceptualization, Supervision, Writing – review & editing

## Acknowledgements

The authors would also like to acknowledge the work undertaken by the Principal Investigators, Site Coordinators and Data Managers involved in the included studies. Most importantly, the authors would like to thank study participants for their contribution to this research.

This research was produced in whole or part by UNSW Sydney researchers and is subject to the UNSW Intellectual property policy. For the purposes of Open Access, the author has applied a Creative Commons Attribution CC BY licence to any Author Accepted Manuscript (AAM) version arising from this submission.

## References

1. Martinello M, Solomon SS, Terrault NA, Dore GJ. Hepatitis C. The Lancet. 2023;402(10407):1085–96.

2. Carson J, Barbieri S, Matthews G, Dore G, Hajarizadeh B. Increasing national trend of direct-acting antiviral discontinuation among people treated for HCV 2016–2021. Hepatol Commun. 2023;7(4).

3. Martinez A, Cheng WH, Marx SE, Manthena S, Dylla DE, Wilson L, et al. Shorter Duration Hepatitis C Virus Treatment is Associated with Better Persistence to Prescription Refills in People Who Inject Drugs: A Real-World Study. Adv Ther. 2023 Aug;40(8):3465–77.

4. Beiser ME, Shaw LC, Shores SK, Carson JM, Hajarizadeh B. Hepatitis C Virus Reinfection in a Real-world Cohort of Homeless-experienced Individuals in Boston. Clin Infect Dis. 2023 Mar;77(1):46–55.

5. Heo M, Norton BL, Pericot-Valverde I, Mehta SH, Tsui JI, Taylor LE, et al. Optimal hepatitis C treatment adherence patterns and sustained virologic response among people who inject drugs: The HERO study. J Hepatol. 2024;80(5):702–13.

6. Chan J, Akiyama MJ, Julian E, Joseph R, McGahee W, Rosner Z, et al. Treating Hepatitis C Virus Infection in Jails as an Offset to Declines in Treatment Activity in the Community, New York City, NY, 2014–2020. AJPM Focus. 2024 Apr 1;3(2):100185.

7. Churkin A, Kriss S, Uziel A, Goyal A, Zakh R, Cotler SJ, et al. Machine learning for mathematical models of HCV kinetics during antiviral therapy. Math Biosci. 2022 Jan 1;343:108756.

8. Goyal A, Lurie Y, Meissner EG, Major M, Sansone N, Uprichard SL, et al. Modeling HCV cure after an ultra-short duration of therapy with direct acting agents. Antiviral Res. 2017;144:281–5.

9. Chua JV, Ntem-Mensah A, Abutaleb A, Husson J, Mutumbi L, Lam KW, et al. Short-duration treatment with the novel non-nucleoside inhibitor CDI-31244 plus sofosbuvir/velpatasvir for chronic hepatitis C: An openlabel study. J Med Virol. 2021 Jun;93(6):3752–60.

10. Sulkowski MS, Flamm S, Kayali Z, Lawitz EJ, Kwo P, McPhee F, et al. Short‐duration treatment for chronic hepatitis C virus with daclatasvir, asunaprevir, beclabuvir and sofosbuvir (FOUR ward study). Liver Int. 2017 Jun;37(6):836–42.

11. Etzion O, Dahari H, Yardeni D, Issachar A, Nevo-Shor A, Cohen-Naftaly M, et al. Response guided therapy for reducing duration of direct acting antivirals in chronic hepatitis C infected patients: a Pilot study. Sci Rep. 2020 Oct 20;10(1):17820.

12. Flower B, Hung LM, Mccabe L, Ansari MA, Le Ngoc C, Vo Thi T, et al. Efficacy of ultra-short, response-guided sofosbuvir and daclatasvir therapy for hepatitis C in a single-arm mechanistic pilot study. eLife. 2023 Jan 9;12:e81801.

13. Canini L, Imamura M, Kawakami Y, Uprichard SL, Cotler SJ, Dahari H, et al. HCV kinetic and modeling analyses project shorter durations to cure under combined therapy with daclatasvir and asunaprevir in chronic HCV-infected patients. PloS One. 2017;12(12):e0187409.

14. Dasgupta S, Imamura M, Gorstein E, Nakahara T, Tsuge M, Churkin A, et al. Modeling-Based Response-Guided Glecaprevir-Pibrentasvir Therapy for Chronic Hepatitis C to Identify Patients for Ultrashort Treatment Duration. J Infect Dis. 2020 Sep 1;222(7):1165–9.

15. Dahari H, Canini L, Graw F, Uprichard SL, Araújo ESA, Penaranda G, et al. HCV kinetic and modeling analyses indicate similar time to cure among sofosbuvir combination regimens with daclatasvir, simeprevir or ledipasvir. J Hepatol. 2016 Jun 1;64(6):1232–9.

16. Lau G, Benhamou Y, Chen G, Li J, Shao Q, Ji D, et al. Efficacy and safety of 3-week response-guided triple direct-acting antiviral therapy for chronic hepatitis C infection: a phase 2, open-label, proof-of-concept study. Lancet Gastroenterol Hepatol. 2016;1(2):97–104.

17. Matthews GV, Bhagani S, Van der Valk M, Rockstroh J, Feld JJ, Rauch A, et al. Sofosbuvir/velpatasvir for 12 vs. 6 weeks for the treatment of recently acquired hepatitis C infection. J Hepatol. 2021 Oct 1;75(4):829–39.

18. Martinello M, Bhagani S, Gane E, Orkin C, Cooke G, Dore GJ, et al. Shortened therapy of eight weeks with paritaprevir/ritonavir/ombitasvir and dasabuvir is highly effective in people with recent HCV genotype 1 infection. J Viral Hepat. 2018 Oct;25(10):1180–8.

19. Martinello M, Orkin C, Cooke G, Bhagani S, Gane E, Kulasegaram R, et al. Short-Duration Pan-Genotypic Therapy With Glecaprevir/Pibrentasvir for 6 Weeks Among People With Recent Hepatitis C Viral Infection. Hepatology. 2020;72(1):7–18.

20. Martinello M, Bhagani S, Shaw D, Orkin C, Cooke G, Gane E, et al. Glecaprevir-pibrentasvir for 4 weeks among people with recent HCV infection: The TARGET3D study. JHEP Rep. 2023 Oct 1;5(10):100867.

21. Yee J, Carson JM, Hajarizadeh B, Hanson J, O’Beirne J, Iser D, et al. High Effectiveness of Broad Access Direct-Acting Antiviral Therapy for Hepatitis C in an Australian Real-World Cohort: The REACH-C Study. Hepatol Commun. 2022;6(3):496–512.

22. Martinello M, Carson JM, Post JJ, Finlayson R, Baker D, Read P, et al. Control and Elimination of Hepatitis C Virus Among People With HIV in Australia: Extended Follow-up of the CEASE Cohort (2014–2023). Open Forum Infect Dis. 2024 Dec 1;11(12):ofae665.

23. Hajarizadeh B, Carson JM, Byrne M, Grebely J, Cunningham E, Amin J, et al. Incidence of hepatitis C virus infection in the prison setting: The SToP-C study. J Viral Hepat [Internet]. 2023 Nov 7 [cited 2023 Dec 1];n/a(n/a). Available from: 10.1111/jvh.13895

24. Dore GJ, Feld JJ, Thompson A, Martinello M, Muir AJ, Agarwal K, et al. Simplified monitoring for hepatitis C virus treatment with glecaprevir plus pibrentasvir, a randomised non-inferiority trial. J Hepatol. 2020 Mar;72(3):431–40.

25. Smolders EJ, Jansen AME, ter Horst PGJ, Rockstroh J, Back DJ, Burger DM. Viral Hepatitis C Therapy: Pharmacokinetic and Pharmacodynamic Considerations: A 2019 Update. Clin Pharmacokinet. 2019;58(10):1237–63.

26. Botros M, Sikaris KA. The De Ritis Ratio: The Test of Time. Clin Biochem Rev. 2013 Nov;34(3):117.

27. Kernbach JM, Staartjes VE. Machine learning-based clinical prediction modeling -- A practical guide for clinicians [Internet]. arXiv; 2020 [cited 2025 Feb 3]. Available from: http://arxiv.org/abs/2006.15069

28. Cawley GC, Talbot NLC. On Over-fitting in Model Selection and Subsequent Selection Bias in Performance Evaluation. J Mach Learn Res. 2010;11(70):2079–107.

29. Japkowicz N, Stephen S. The class imbalance problem: A systematic study. Intell Data Anal. 2002 Jan 1;6(5):429–49.

30. Han H, Wang WY, Mao BH. Borderline-SMOTE: A New Over-Sampling Method in Imbalanced Data Sets Learning. In: Huang DS, Zhang XP, Huang GB, editors. Advances in Intelligent Computing. Berlin, Heidelberg: Springer; 2005. p. 878–87.

31. Collins GS, Moons KGM, Dhiman P, Riley RD, Beam AL, Calster BV, et al. TRIPOD+AI statement: updated guidance for reporting clinical prediction models that use regression or machine learning methods. BMJ. 2024 Apr 16;385:e078378.

32. Park H, Lo-Ciganic WH, Huang J, Wu Y, Henry L, Peter J, et al. Evaluation of machine learning algorithms for predicting direct-acting antiviral treatment failure among patients with chronic hepatitis C infection. Sci Rep. 2022;12(1):18094.

33. Medeiros-Filho JE, e Guedes de Carvalho Mello IMV, Rebello Pinho JR, Neumann AU, de Mello Malta F, Caetano da Silva L, et al. Differences in viral kinetics between genotypes 1 and 3 of hepatitis C virus and between cirrhotic and non-cirrhotic patients during antiviral therapy. World J Gastroenterol. 2006 Dec 7;12(45):7271–7.

34. Ward JC, Bowyer S, Chen S, Fernandes Campos GR, Ramirez S, Bukh J, et al. Insights into the unique characteristics of hepatitis C virus genotype 3 revealed by development of a robust sub-genomic DBN3a replicon. J Gen Virol. 2020 Nov;101(11):1182–90.

35. Thomas DL, Owen A, Kiser JJ. Prospects for Long-Acting Treatments for Hepatitis C. Clin Infect Dis. 2022 Dec 1;75(Supplement_4):S525–9.

36. Carson JM, Dore GJ. Learning From the Gaps: Rethinking Hepatitis C Virus Retreatment for People Who Inject Drugs. Clin Infect Dis. 2025 Apr 15;ciaf083.

37. Han R, Acosta JN, Shakeri Z, Ioannidis JPA, Topol EJ, Rajpurkar P. Randomised controlled trials evaluating artificial intelligence in clinical practice: a scoping review. Lancet Digit Health. 2024 May 1;6(5):e367–73.

38. Tonekaboni S, Joshi S, McCradden MD, Goldenberg A. What Clinicians Want: Contextualizing Explainable Machine Learning for Clinical End Use. In: Proceedings of the 4th Machine Learning for Healthcare Conference [Internet]. PMLR; 2019 [cited 2024 Nov 5]. p. 359–80. Available from: https://proceedings.mlr.press/v106/tonekaboni19a.html

39. Welzel TM, Yang M, Sajeev G, Chen YJ, Pinsky B, Bao Y, et al. Assessing Patient Preferences for Treatment Decisions for New Direct Acting Antiviral (DAA) Therapies for Chronic Hepatitis C Virus Infections. Adv Ther. 2019 Sep 1;36(9):2475–86.

40. Slevin AR, Hart MJ, Van Horn C, Rahman S, Samji NS, Szabo A, et al. Hepatitis C virus direct-acting antiviral nonadherence: relationship to sustained virologic response and identification of at-risk patients. J Am Pharm Assoc. 2019;59(1):51–6.

41. Feld JJ, Hoofnagle JH. Mechanism of action of interferon and ribavirin in treatment of hepatitis C. Nature. 2005 Aug;436(7053):967–72.

