## Supplementary Material for "Development of a machine learning model to predict short duration HCV treatment response"

---

### Data privacy and study ethics

The training datasets used in this study are held securely at the Kirby Institute, UNSW Sydney. Due to the sensitive nature of this data, it cannot be made publicly available however, data used in this analysis may be made available for research purposes upon reasonable request to the corresponding author.

All studies contributing data to this analysis were conducted according to the Declaration of Helsinki and International Conference on Harmonization Good Clinical Practice (ICH/GCP) guidelines. Study details and ethics approvals can be found below:

Randomised study of interferon-free treatment for recently acquired HCV in people who inject drugs and people with HIV coinfection (**REACT**(1)) enrolled participants (n= at 24 primary and tertiary sites in Australia (n=5), Canada (n=4), Germany (n=4), Netherlands (n=1), New Zealand (n=1), Switzerland (n=3), United Kingdom (n=4), and United States (n=2) (1). Ethics: Royal Adelaide Hospital Human Research Ethics Committee (Australia), as well as through local ethics committees at all study sites. Study was conducted March 2017 to December 2019. Clinical Trial Registration: NCT02625909.

---

Treatment of recently acquired hepatitis C with the 3D regimen or G/P (**TARGET3D**) was conducted in 3 phases:

i. Treatment with paritaprevir-ritonavir-ombitasvir+dasabuvir 8-weeks enrolled participants (n=30) at 6 tertiary sites in Australia (n=1), England (n=4) and New Zealand (n=1) (2). Study was conducted June 2016 to August 2017.
ii. Treatment with glecaprevir-pibrentasvir 6-weeks enrolled participants (n=30) at 8 tertiary sites in Australia (n=2), England (n=5), and New Zealand (n=1) (3). Study was conducted October 2018 to November 2018.
iii. Treatment with glecaprevir-pibrentasvir 4-weeks enrolled participants (n=30) at 9 tertiary hospital clinics in Australia (n = 3), England (n = 5) and New Zealand (n = 1) (4). Study was conducted December 2018 to April 2021.

Ethics: St Vincent's Hospital, Sydney Human Research Ethics Committee (Australia), Health and Disabilities Ethics Committee (New Zealand), London-Riverside Research Ethics Committee (England), and local ethics and governance committees at all sites. Clinical Trial Registration: NCT02634008.

---

Strategic treatment reduction in very early liver disease (**STRIVE4**) enrolled participants (n=4; recruitment ongoing) at 4 tertiary sites in Australia. Ethics Approval: St Vincent's Hospital, Sydney Human Research Ethics Committee (Australia) and local ethics committees. Study commenced in January 2020 and is ongoing (no publication for reference available at this time). Clinical Trial Registration: NCT03855917.

---

A phase IIIb, open-label, multicentre, international randomised controlled trial of simplified treatment monitoring for 8 weeks glecaprevir-pibrentasvir in chronic HCV treatment naïve patients without cirrhosis (**SMART-C** (5)). Enrolled participants (n=380) at 33 primary and tertiary sites in Australia (n=6), Canada (n=7), France (n = 3), Germany (n=4), New Zealand (n=4), Switzerland (n=2), the United Kingdom (n=3), and the United States (n=4). Study was conducted August 2017 to November 2018. Ethics: St Vincent's Hospital Human Research Ethics (Australia), Western Institutional Review Board (Canada), Veritas Independent Review Board (Canada), Willian Osler Health Service Research Ethics Board (Canada), Hamilton Integrated Research Ethics Board (Canada), Comité d'éthique de la recherche CHU de Québec - Université Laval (Canada), Comité de Protection des Personnes (France), Ethikkommision Medizinischen Hochshule Hannover (Germany), Health and Disability Ethics Committee (New Zealand), Schweizerische Ethikkommissionen für die Forschung am Menschen (Switzerland), A Research Ethics Committee established by the Health Research Authority, London - South East Research Ethics Committee (United Kingdom), Beth Isreal Deaconess Medical Centre Committee on Clinical Investigations (United States), Duke Medicine Institutional Review Board for Clinical Investigations (United States), New York University School of Medicine Institutional Review Board (United States), Dean Institutional Review Board (United States). Trial Registration: NCT03117569

---

Control and Elimination of HCV among people with HIV (**CEASE**) clinical cohort study enrolled participants (n=402) at 14 primary and tertiary sites in Australia (6). Ethics: St Vincent's Hospital, Sydney Human Research Ethics Committee (Australia), and local governance committees at all sites. Study was conducted July 2014 to February 2023. Clinical Trial Registration: NCT02102451

---

Surveillance and Treatment of People in Prison with HCV (**STOP-C**) enrolled participants (n=3691) at 4 prison sites in Australia (7). Ethics: New South Wales Justice Health and Forensic Mental Health Network Human Research Ethics Committee (Australia), Aboriginal Health and Medical Research Council Human Research Ethics Committee (Australia), and New South Wales Corrective Services Ethics Committee (Australia). Study was conducted October 2014 to December 2019. Clinical Trial Registration: NCT02064049.

---

Real world effectiveness of antiviral therapy for HCV (**REACH-C**) standard-of-care cohort extracted data for individuals (n=10,834) from 33 primary and tertiary health services across Australia (8). Ethics: St Vincent's Hospital, Sydney Human Research Ethics Committee (Australia), Aboriginal Health and Medical Research Council (Australia), Northern Territory Human Research Ethics Committee (Australia), Central Australian Human Research Ethics Committee (Australia), Western Australian Aboriginal Health Ethics Committee (Australia) and Tasmanian Health and Medical Research Ethics Committee (Australia). Study was conducted March 2016 to October 2020. Clinical Trial Registration: NA

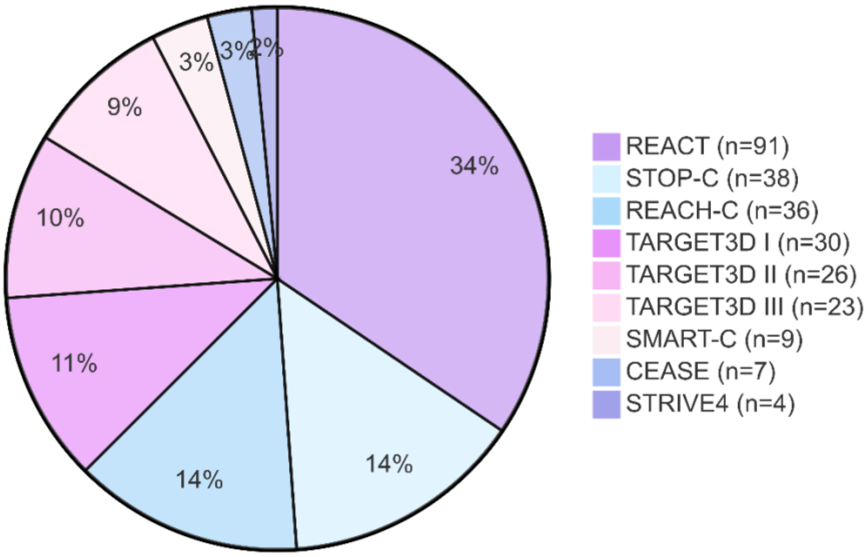

**Supplementary Figure 1. Proportional contributions of data sources.**

Pie chart illustrates the proportional contributions of data from nine studies that comprise the total dataset (n=264). The data sources from clinical trials (n=183; REACT; TARGETED; SMART-C; STRIVE4) and cohort studies (n=81; STOP-C; REACH-C; CEASE).

### Performance metrics

From the testing data, the number of true positives (TP; both the classifier and data indicate the target outcome), true negatives (TN; both the classifier and data did not indicate the target outcome), false positives (FP: classifier predicted the target outcome, but the data disagree) and false negatives (FN; classifier does not predict the target outcome, but the data disagree) were identified. The below performance metrics were calculated with treatment failure as the target (or positive) outcome:

**Sensitivity (True Positive Rate, Recall)** is the proportion of actual positive cases correctly identified by the model. It measures the ability of the model to detect positive cases.

Sensitivity (TPR) = TP / (TP + FN)

Sensitivity ranges from 0 to 1 (or 0 to 100%). Higher sensitivity indicates fewer false negatives, making it critical in contexts where missing positive cases is costly.

---

**Specificity (True Negative Rate)** is the proportion of actual negative cases correctly identified by the model. It measures the ability of the model to avoid false positives.

Specificity (TNR) = TN / (TN + FP)

Specificity ranges from 0 to 1 (or 0 to 100%). Higher specificity indicates fewer false positives, crucial in scenarios where false alarms should be minimized.

---

**Positive Predictive Value (PPV, Precision)** measures the proportion of predicted positive cases that are true positives. It reflects the reliability of positive predictions.

PPV = TP / (TP + FP)

PPV ranges from 0 to 1 (or 0 to 100%). High PPV indicates that a positive prediction is likely to be correct, important in clinical contexts to avoid unnecessary treatments.

---

**Negative Predictive Value (NPV)** measures the proportion of predicted negative cases that are true negatives. It reflects the reliability of negative predictions.

NPV = TN / (TN + FN)

NPV ranges from 0 to 1 (or 0 to 100%). High NPV indicates that a negative prediction is likely to be correct, critical when ruling out a condition or disease.

---

**Area Under the Receiver Operating Characteristic Curve (AUROC)** is a measure of a model's ability to distinguish between positive and negative classes across all possible classification thresholds. AUROC plots the True Positive Rate (Sensitivity) against the False Positive Rate (1 - Specificity). While AUROC is computed numerically, it is conceptually:

$$\text{AUROC} = \int_0^1 \text{Sensitivity}\ d(1 - \text{Specificity})$$

AUROC ranges from 0.5 to 1; where scores of 0.5 indicate no discrimination (random guessing) and scores of 1 indicate perfect discrimination.

---

**Area Under the Precision-Recall Curve (AUPRC)** quantifies the relationship between Precision (PPV) and Recall (Sensitivity) across all possible classification thresholds. It is particularly valuable when dealing with imbalanced datasets, where the positive class is underrepresented. While AUPRC is computed numerically, it represents:

AUPRC= $\int$ Precision $d$(Recall)

AUPRC ranges 0 to 1; where scores of 0 indicate no meaningful predictions and scores of 1 indicate perfect predictions.

---

**The Fowlkes-Mallows Index (FMI)** is the geometric mean of precision (PPV) and recall (sensitivity; TPR) that balances the two metrics. It is useful in evaluating binary classification tasks, when the consequences of false negatives and false positives need careful consideration.

FMI = $\sqrt{}$ (PPV * TPR)

FMI ranges from 0 to 1; where scores of 0 indicates poor performance in accurately predicting true positives, and a score of 1 indicates perfection predictions.

---

**Matthews Correlation Coefficient (MCC)** provides a balanced measure of the quality of binary classifications, accounting for all four confusion matrix categories (TP, TN, FP, FN). It is especially useful when the dataset is imbalanced.

MCC = (TP * TN – FP * FN) / sqrt ((TP + FP) * (TP + FN) * (TN + FP) * (TN + FN))

MCC ranges from -1 to +1; score of +1 indicate perfect classification, scores of 0 indicate no better than random guessing, scores of -1 indicate completely incorrect classification.

---

**Youden Index** is a summary measure used to identify the optimal classification threshold that maximized the balance between sensitivity and specificity. It is defined as:

Youden Index = Sensitivity + Specificity−1

The Youden Index ranges from -1 to 1, with 1 indicating perfect classification performance and 0 representing a threshold with no discriminatory power.

---

**Quadratic Cost Function** penalizes classification errors based on the confidence of predictions, ensuring that highly confident errors are penalized more heavily than less confident ones. The total cost for a given threshold is computed as:

$$\text{Total Cost} = w_{\text{FP}} \sum_{i \in \text{FP}} (1 - y_{\text{proba}}[i])^2 + w_{\text{FN}} \sum_{i \in \text{FN}} y_{\text{proba}}[i]^2 + w_{\text{TP}} \sum_{i \in \text{TP}} (1 - y_{\text{proba}}[i])^2$$

This approach is particularly suited for scenarios where incorrect classifications carry varying clinical or economic consequences. For example, when assignment of incorrect treatment durations results in excess treatment or retreatment costs.

**Supplementary Table 1: TRIPOD checklist.**

Transparent Reporting of a Multivariable Prediction Model for Individual Prognosis or Diagnosis (TRIPOD) (9)

| Section/Topic | Item | Checklist Item | Section |
|---|---|---|---|
| Title | 1 | Identify the study as developing and/or validating a multivariable prediction model, the target population, and the outcome to be predicted. | T |
| Abstract | 2 | Provide a summary of objectives, study design, setting, participants, sample size, predictors, outcome, statistical analysis, results, and conclusions. | A |
| Background & Objectives | 3a | Explain the medical context (including whether diagnostic or prognostic) and rationale for developing or validating the multivariable prediction model, including references to existing models. | A, I |
| | 3b | Specify the objectives, including whether the study describes the development or validation of the model or both. | I, M |
| Source of data | 4a | Describe the study design or source of data (e.g., randomized trial, cohort, or registry data), separately for the development and validation data sets, if applicable. | M, S |
| | 4b | Specify the key study dates, including start of accrual; end of accrual; and, if applicable, end of follow-up. | NA |
| Participants | 5a | Specify key elements of the study setting (e.g., primary care, secondary care, general population) including number and location of centers. | R, S |
| | 5b | Describe eligibility criteria for participants. | M |
| | 5c | Give details of treatments received, if relevant. | M, R |
| Outcome | 6a | Clearly define the outcome that is predicted by the prediction model, including how and when assessed. | I, M |
| | 6b | Report any actions to blind assessment of outcome to be predicted | M |
| Predictors | 7a | Clearly define all predictors used in developing or validating the multivariable prediction model, including how and when they were measured. | M, R |
| | 7b | Report any actions to blind assessment of predictors for the outcome and other predictors. | NA |
| Sample Size | 8 | Explain how the study sample size was arrived at. | M |
| Missing Data | 9 | Describe how missing data were handled (e.g., complete-case analysis, single imputation, multiple imputation) with details of any imputation method. | M |
| Statistical Analysis Methods | 10a | Describe how predictors were handled in the analyses | M, R |
| | 10b | Specify type of model, all model-building procedures (including any predictor selection), and method for internal validation. | M, R |
| | 10d | Specify all measures used to assess model performance and, if relevant, to compare multiple models. | M, S |
| Risk Groups | 11 | Provide details on how risk groups were created, if done. | NA |
| Participants | 13a | Describe the flow of participants through the study, including the number of participants with and without the outcome and, if applicable, a summary of the follow-up time. | R |
| | 13b | Describe the characteristics of the participants (basic demographics, clinical features, available predictors) | R, T1 |
| Model Development | 14a | Specify the number of participants and outcome events in each analysis | R, T1 |
| | 14b | If done, report the unadjusted associated between each candidate predictor and outcome | NA |
| Model Specification | 15a | Present the full prediction model to allow predictions for individuals (ie, all regression coefficients and model intercept or baseline survival at a given point) | S |
| | 15b | Explain how to use the prediction model | NA |
| Model performance | 16 | Report performance measures for the prediction model | R |
| Limitations | 18 | Discuss any limitations of the study (such as non-representative sample, few events per predictor, missing data) | D |
| Interpretation | 19 | Give and overall interpretation of the results, considering objectives, limitations and results from similar studies | D |
| Implications | 20 | Discuss the potential clinical use of the model and implications for future research | D |
| Supplementary Material | 21 | Provide information about the availability of supplementary resources such as the study protocol, web calculator and data sets | S |
| Funding | 22 | Give the source of funding and the role of the funders in the present study | S |

**Abbreviations:** T, title; A, abstract; I, introduction; M, methods; R, results; D, discussion, S, supplement; T, table; ST, supplementary table; F, figure; SF, supplementary figure; NA, not applicable.

**Supplementary Figure 2. Stages of ML model development and application.**
Flow chart diagramming the key phases of machine learning model development - training, validation, and deployment - alongside example clinical scenarios that highlight potential use cases.

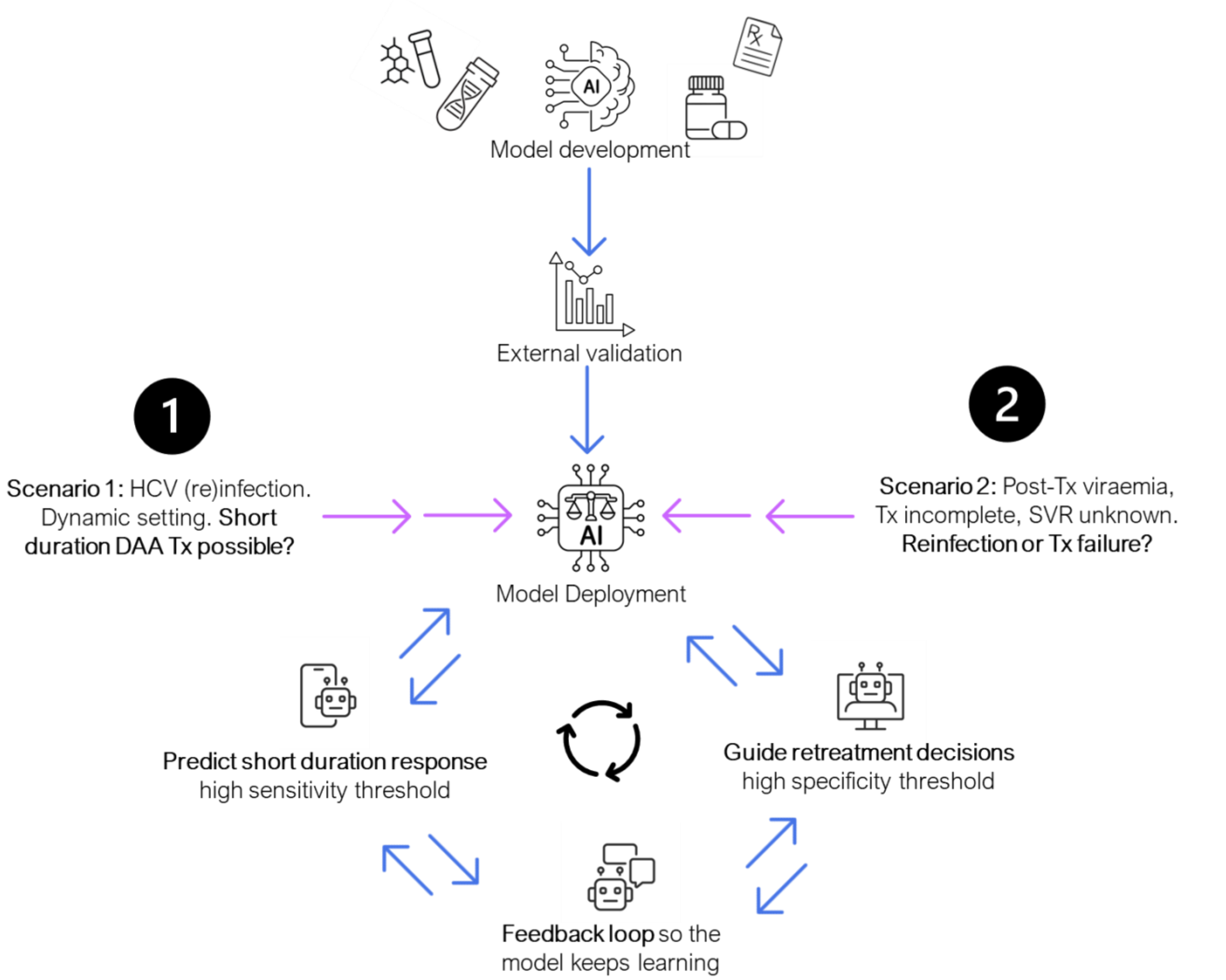

**Abbreviations:** ML, machine learning; Tx, treatment; SVR, sustained virologic response

**Supplementary Figure 3. Violin plots comparing measures for acute vs. chronic HCV.**
Violin plots illustrate the distribution of HCV RNA levels and liver function markers (e.g., AST-ALT ratio, APRI, ALT, and AST) stratified by HCV phase (chronic vs. acute). Median values and variability within each group are shown, with p-values calculated using the Mann-Whitney U test to assess differences between groups.

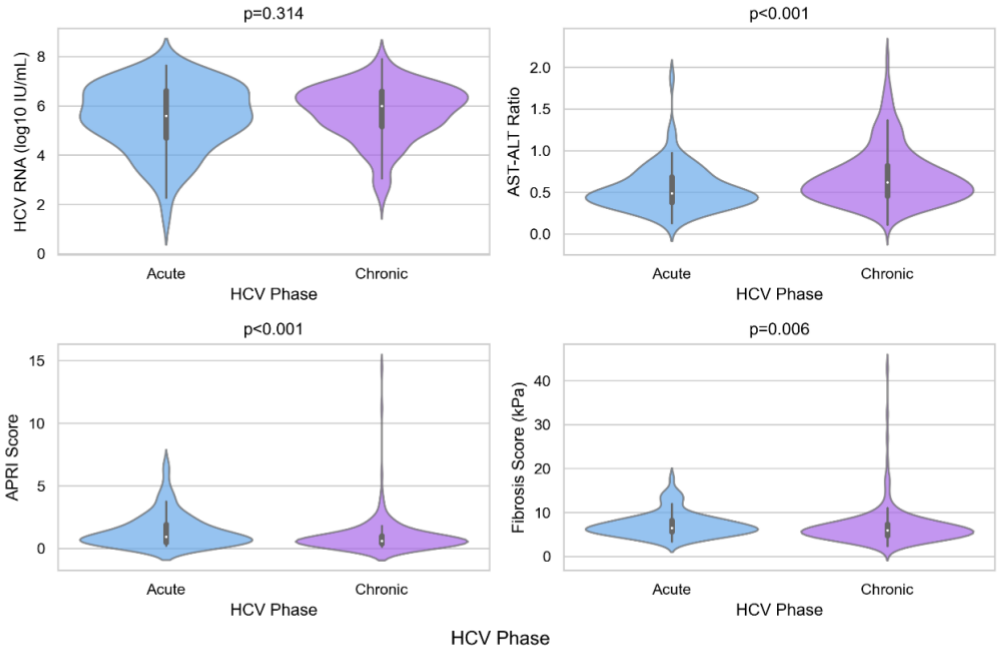

**Supplementary Figure 4. Scatter plots of correlation between measures of liver function.**
Scatter plots depict correlations between liver function markers, with Pearson correlation coefficients (r) to quantify strength and direction of linear relationships (r=-1 indicates perfect negative correlation; r=0 indicates no correlation; r=1 indicates perfect positive correlation). Data points are color-coded by treatment response: blue dots represent sustained virological response, and purple dots represent treatment failure.

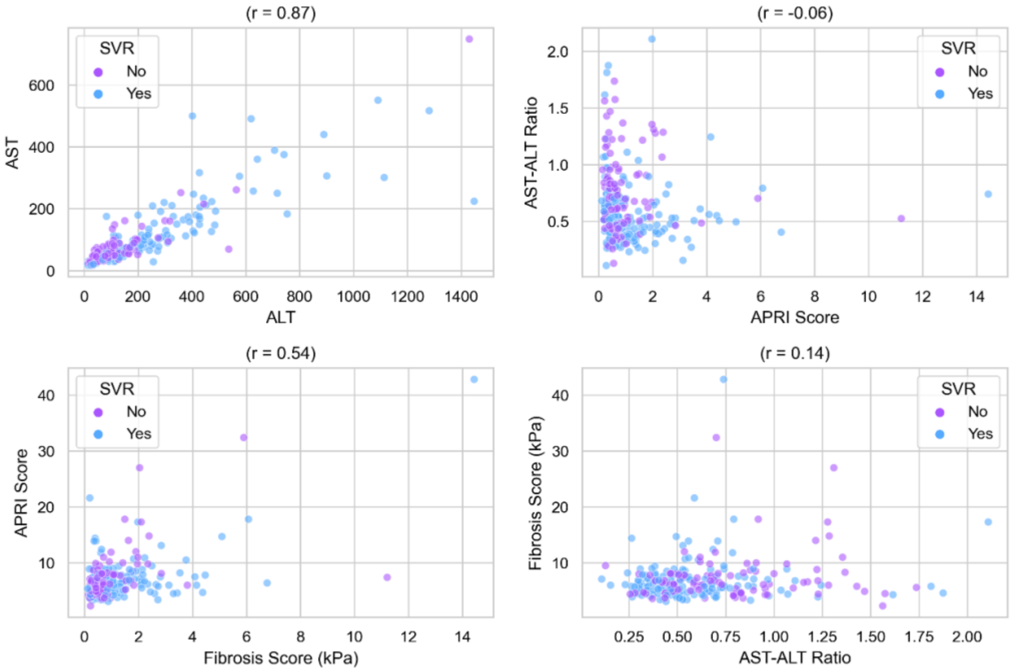

**Abbreviations:** HCV, hepatitis C virus; ALT, alanine aminotransferase; AST, aspartate aminotransferase; Fibrosis Score, liver stiffness measure; APRI, aspartate aminotransferase-to-platelet ratio index.

**Supplementary Table 2. Clinical trial and cohort populations.**

Comparison of the clinical trial and cohort populations, reporting p-values to assess statistical significance. The clinical trial population predominantly comprised individuals treated with short-duration direct-acting antivirals to assess efficacy, whereas the cohort study population largely included individuals who discontinued standard-duration therapy.

| | Clinical Trial (n=183) | Cohort (n=81) | p-value |
|---|---|---|---|
| Age, median [Q1, Q3] | 45.7 [36.2,53.5] | 35.2 [27.4,50.0] | <0.001 |
| Male gender, n (%) | 174 (95.1) | 71 (87.7) | 0.058 |
| Caucasian/white race, n (%) | 137 (75.6) | 76 (94.3) | <0.001 |
| Gay or bisexual man, n (%) | 146 (79.8) | 7 (8.6) | <0.001 |
| IDU past six months, n (%) | 72 (39.3) | 41 (50.6) | 0.116 |
| Incarcerated, n (%) | 0 (0.0) | 38 (46.9) | <0.001 |
| HIV, n (%) | 123 (67.2) | 8 (9.9) | <0.001 |
| Acute HCV, n (%) | 83 (45.4) | 1 (1.2) | <0.001 |
| HCV RNA $log_{10}$ IU/L, median [Q1, Q3] | 5.8 [5.1,6.6] | 6.1 [5.0,6.6] | 0.579 |
| Genotype, n (%) | | | |
| 1 | 133 (72.7) | 34 (42.0) | <0.001 |
| 3 | 22 (12.0) | 45 (55.6) | |
| 2, 4, 6 | 28 (15.3) | 2 (2.4) | |
| AST-ALT ratio, median [Q1, Q3] | 0.5 [0.4,0.7] | 0.6 [0.5,0.9] | 0.221 |
| APRI, median [Q1, Q3] | 0.7 [0.4,1.5] | 0.6 [0.4,1.0] | 0.311 |
| Liver stiffness (kPa), median [Q1, Q3] | 6.0 [4.8,7.5] | 6.2 [5.0,8.0] | 0.195 |
| Cirrhosis, n (%) | 7 (3.8) | 8 (9.9) | 0.089 |
| DAA classes, n (%) | | | |
| NS5A+NS5B | 91 (49.7) | 71 (87.7) | <0.001 |
| PI+NS5A | 61 (33.3) | 9 (11.1) | |
| PI+NS5A+NS5B | 31 (16.9) | 1 (1.2) | |
| Duration (days), median [Q1, Q3] | 42.0 [42.0, 42.0] | 28.0 [27.0, 40.0] | <0.001 |

**Abbreviations:** IDU, injecting drug use; ALT, alanine aminotransferase; AST, aspartate aminotransferase; PLT, platelet count; APRI; AST to Platelet Ratio Index, DAA, direct acting antiviral; PI; protease inhibitor.

**Supplementary Table 3. Logistic regression analysis.**

Logistic regression analysis identifying factors associated with short-duration treatment failure. Results are presented as adjusted odds ratios with 95% confidence intervals and p-values to quantify the strength and significance of associations.

| | Adjusted Odds Ratio | 95% CI | p-value |
|---|---|---|---|
| Age (years) | 0.99 | 0.96-1.02 | 0.446 |
| Gender | | | |
| Female | 1.00 | - | - |
| Male | 0.71 | 0.19-2.54 | 0.594 |
| Race | | | |
| Other | 1.00 | - | - |
| Caucasian/white | 1.01 | 0.36-2.81 | 0.980 |
| IDU (last six months) | | | |
| No | 1.00 | - | - |
| Yes | 0.50 | 0.25-1.00 | 0.070 |
| HCV RNA ($log_{10}$ IU/L) | 2.10 | 1.51-2.92 | <0.001 |
| Genotype | | | |
| Non-genotype 3 | 1.00 | - | - |
| Genotype 3 | 3.16 | 1.52-6.61 | 0.002 |
| AST-ALT ratio | 7.48 | 2.50-22.39 | <0.001 |
| APRI | 0.81 | 0.61-1.07 | 0.135 |
| Liver fibrosis (kPa) | 1.02 | 0.91-1.14 | 0.759 |
| DAA class | | | |
| NS5A+NS5B | 1.00 | - | - |
| PI+NS5A | 0.31 | 0.14-0.68 | 0.004 |
| PI+NS5A+NS5B | 0.36 | 0.07-2.04 | 0.251 |
| DAA duration (days) | 0.90 | 0.86-0.94 | <0.001 |

**Abbreviations:** DAA, direct acting antiviral; HCV, hepatitis C virus; ALT, alanine aminotransferase; AST, aspartate aminotransferase; Fibrosis, liver stiffness measured by transient elastography (kPa); APRI, aspartate aminotransferase-to-platelet ratio index; IDU, injecting drug use past six months; PI, protease inhibitor.

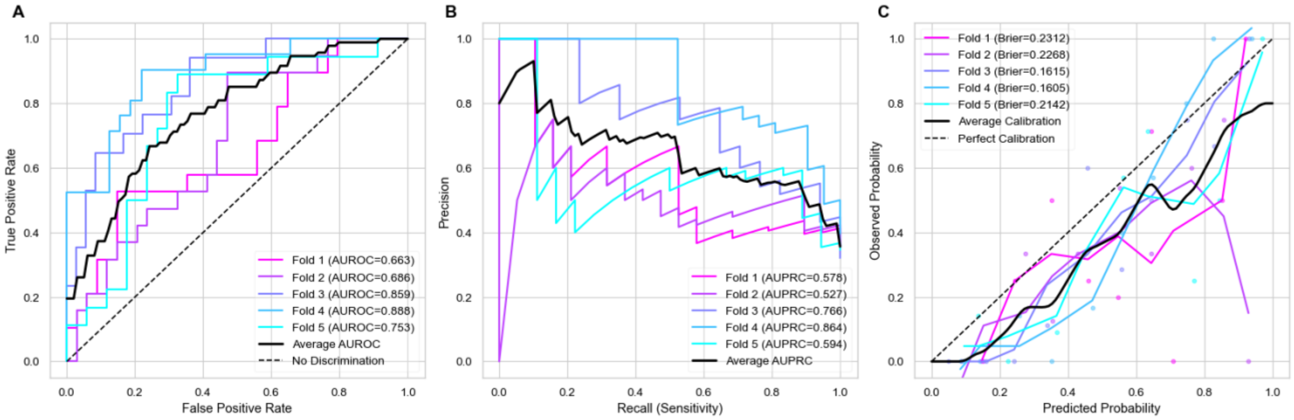

**Supplementary Figure 5. Nested cross-validation for logistic regression.**

Evaluation of the logistic regression model's predictive accuracy through nested cross-validation, presented with **(A)** AUROC (Area Under the Receiver Operating Characteristic Curve), **(B)** AUPRC (Area Under the Precision-Recall Curve), and **(C)** Brier Calibration Curve to provide insight with Savitzky–Golay smoothing as to how well the model generalizes across varying subsets of data for predicting HCV treatment response.

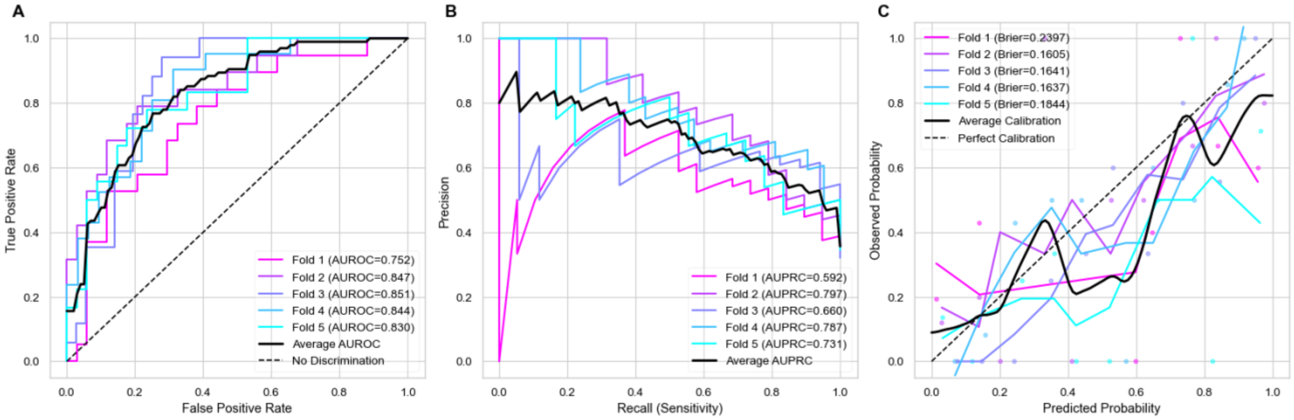

**Supplementary Figure 6. Nested cross-validation for XGBoost.**

Evaluation of the XGBoost model's predictive accuracy through nested cross-validation, presented with **(A)** AUROC (Area Under the Receiver Operating Characteristic Curve), **(B)** AUPRC (Area Under the Precision-Recall Curve), and **(C)** Brier Calibration Curve to provide insight with Savitzky–Golay smoothing as to how well the model generalizes across varying subsets of data for predicting HCV treatment response.

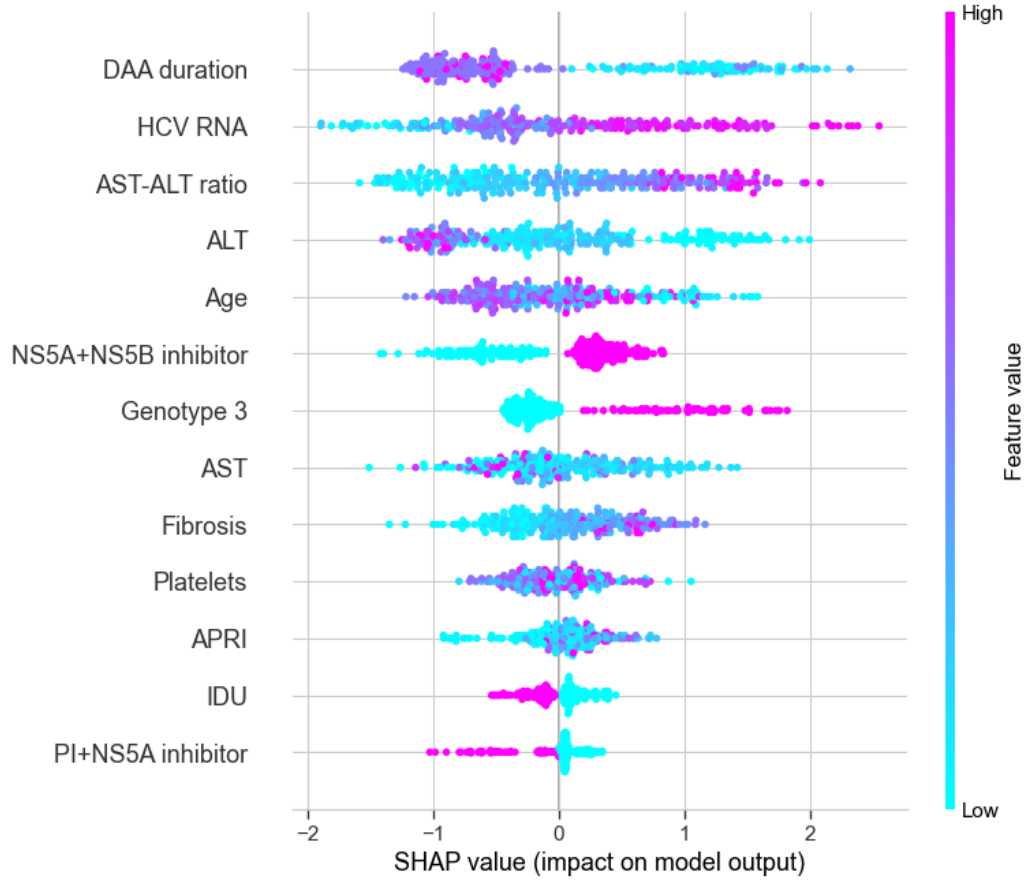

**Supplementary Figure 7. SHAP analysis for the XGBoost model.**
Feature Importance in Predicting HCV Treatment Failure. SHAP (SHapley Additive exPlanations) plot illustrating feature contributions to the Elastic Net model's predictions of treatment failure in HCV patients. Each point represents a SHAP value for a feature in a given observation, where colour indicates feature value (e.g., high or low). Positive SHAP values suggest a higher probability of treatment failure, while negative values suggest a greater likelihood of cure.

**Abbreviations:** DAA, direct acting antiviral; HCV, hepatitis C virus; ALT, alanine aminotransferase; AST, aspartate aminotransferase; Fibrosis, liver stiffness measured by transient elastography (kPa); APRI, aspartate aminotransferase-to-platelet ratio index; IDU, injecting drug use past six months; PI, protease inhibitor.

### Final model parameters

**Logistic Regression**
LogisticRegression(C=1.0, solver: 'lbfgs', penalty: ‘none’, class_weight: None, max_iter=100, random_state=23)

**Elastic Net**
LogisticRegression(C=0.615848211066026, class_weight='balanced', l1_ratio=0.306, max_iter=500, penalty='elasticnet', random_state=23, solver='saga',  tol=1e-06)

**XGBoost**
XGBClassifier(colsample_bytree: 0.8, subsample: 0.8, learning_rate: 0.1, max_depth: 3, n_estimators: 300, objective: binary:logistic, random_state: 23, 'reg_alpha': 0, reg_lambda: 1)

### References

1. Matthews G, Bhagani S, van der Valk M, Rockstroh J, Feld J, Rauch A, et al. Sofosbuvir/velpatasvir for 12 vs. 6 weeks for the treatment of recently acquired hepatitis C infection. J Hepatol. 2021;75(4):829–39.

2. Martinello M, Bhagani S, Gane E, Orkin C, Cooke G, Dore GJ, et al. Shortened therapy of eight weeks with paritaprevir/ritonavir/ombitasvir and dasabuvir is highly effective in people with recent HCV genotype 1 infection. J Viral Hepat. 2018;25(10):1180–8.

3. Martinello M, Orkin C, Cooke G, Bhagani S, Gane E, Kulasegaram R, et al. Short-Duration Pan-Genotypic Therapy With Glecaprevir/Pibrentasvir for 6 Weeks Among People With Recent Hepatitis C Viral Infection. Hepatology. 2020 Jul;72(1):7–18.

4. Martinello M, Bhagani S, Shaw D, Orkin C, Cooke G, Gane E, et al. Glecaprevir-pibrentasvir for 4 weeks among people with recent HCV infection: The TARGET3D study. JHEP Rep. 2023 Oct 1;5(10):100867.

5. Dore GJ, Feld JJ, Thompson A, Martinello M, Muir AJ, Agarwal K, et al. Simplified monitoring for hepatitis C virus treatment with glecaprevir plus pibrentasvir, a randomised non-inferiority trial. J Hepatol. 2020;72(3):431–40.

6. Martinello M, Carson JM, Post JJ, Finlayson R, Baker D, Read P, et al. Control and Elimination of Hepatitis C Virus Among People With HIV in Australia: Extended Follow-up of the CEASE Cohort (2014–2023). Open Forum Infect Dis. 2024 Dec 1;11(12):ofae665.

7. Hajarizadeh B GJ Byrne B, Marks P, Amin J, McManus H, Butler T, Cunningham EB, Vickerman P, Martin NK, McHutchison JG, Brainard DM, Treloar C, Chambers GM, Grant L, Mcgrath C, Lloyd AR, Dore G. Evaluation of hepatitis C treatment-as-prevention within an Australian prison prospective cohort: The SToP-C study. Lancet Gastroenterol Hepatol. 2021;6(7):533–46.

8. Yee J, Carson JM, Hajarizadeh B, Hanson J, O'Beirne J, Iser D, et al. High Effectiveness of Broad Access Direct-Acting Antiviral Therapy for Hepatitis C in an Australian Real-World Cohort: The REACH-C Study. Hepatol Commun. 2022;6(3):496–512.

9. Collins GS, Moons KGM, Dhiman P, Riley RD, Beam AL, Calster BV, et al. TRIPOD+AI statement: updated guidance for reporting clinical prediction models that use regression or machine learning methods. BMJ. 2024 Apr 16;385:e078378.
